# Neutralizing antibodies against SARS-CoV-2 variants of concern elicited by the Comirnaty® COVID-19 vaccine in nursing home residents

**DOI:** 10.1101/2021.10.06.21264607

**Authors:** Beatriz Sánchez-Sendra, Eliseo Albert, Joao Zulaica, Ignacio Torres, Estela Giménez, Pilar Botija, María José Beltrán, Celia Rodado, Ron Geller, David Navarro

## Abstract

Immunosenescence may impact the functionality and breadth of vaccine-elicited humoral immune responses. The ability of sera to neutralize the SARS-CoV-2 spike protein (S) from Beta, Gamma, Delta, and Epsilon variants of concern (VOCs) relative to the ancestral Wuhan-Hu-1 strain was compared in Comirnaty® COVID-19-vaccinated elderly nursing home residents (n=30) or younger individuals (n=18) and non-vaccinated individuals who recovered from severe COVID-19 (n=19). In all groups, some participants lacked NtAb against one or more VOCs, mainly the Beta variant (15-20%). Serum NtAb titers were lowest against the Beta variant followed by Gamma, Epsilon, and Delta variants. Fold change reduction in NtAb titers relative to the ancestral strain was greatest for the Beta variant (6.7-18.8) followed by Gamma (3.6-6.2), Epsilon (2.9-5.8), and Delta (3.5-4.3) variants, regardless of the study group considered. In summary, older age, frailty, and concurrence of co-morbidities had no impact on the serum NtAb activity profile against SARS-CoV-2 VOCs.

## INTRODUCTION

Coronaviruses encode a proof-reading mechanism to minimize sequence variation in the long RNA genomes that may compromise their survival in nature [1]. Nevertheless, generation of variants during coronavirus replication in humans unfailingly ensues [2]. Many SARS-CoV-2 variants have emerged since its appearance late in 2019, some of which may pose a threat to pandemic control due to their relative resistance to neutralizing activity of antibodies elicited following natural infection or vaccination with the original SARS-CoV-2 isolate (Wuhan/Hu-1); these variants, termed variants of concern (VOC), incorporate amino acid substitutions or deletions in critical locations within the virus Spike (S) protein [3]. The Comirnaty® mRNA COVID-19 vaccine encodes the ancestral strain S protein in a pre-fusion conformation [4]. Several studies have shown a decreased ability of serum antibodies elicited by this vaccine to neutralize a number of VOCs compared to the original strain, most notably Beta (B.1.351), Gamma (P.1), and more recently Delta (B.1.617.2)[5-12]. Nevertheless, elderly people with frailty and co-morbidities, who are at the highest risk to develop severe forms of COVID-19, were either not included or underrepresented in these studies. Since immunosenescence may be detrimental to the functionality and breadth of vaccine-elicited humoral immune responses [13], profiling of cross-neutralizing activity against VOCs of sera from fully vaccinated elderly people is of major public health interest. In the current work, we address this issue by comparing the neutralizing capacity of sera from Comirnaty® COVID-19-vaccinated nursing home residents with that of sera from younger and seemingly healthy vaccinated individuals and non-vaccinated individuals who recovered from severe COVID-19.

## MATERIAL AND METHODS

### Study groups

In this observational retrospective study, three population groups were considered. Group 1 included 30 nursing home (NH) residents with frailty and one or more co-morbidities (23 females), randomly selected from one NH affiliated to the Clínico-Malvarrosa Health Department, Valencia (Spain), aged a median of 90 years (range 64-100). Twenty-two were SARS-CoV-2 naïve, as judged by a lack of antibodies targeting the SARS-CoV-2 nucleoprotein (N), and 8 had contracted COVID-19 requiring hospitalization prior to receiving the first dose of the Comirnaty® COVID-19 vaccine, as determined by positive RT-PCR results in nasopharyngeal specimens and the detection of SARS-CoV-2 N antibodies. In all cases, the variant Wuhan/D614G was involved, as determined by whole-genome sequencing (not shown). All participants in this group were vaccinated on the same day and the sera were drawn at a median of 35 days after the second vaccine dose.

Group 2 comprised 18 healthy individuals (14 females) aged a median of 47.5 years (range, 26-62). No molecular or serological evidence of prior SARS-CoV-2 infection was available from any of these participants, whose sera were drawn at a median of 15 days since the completion of the vaccination schedule (range, 13-18).

Group 3 included 19 individuals (7 females; median age, 63 years; range 40-80) who were infected with the Wuhan/D614G strain (determined by whole-genome sequencing; data not shown), required hospitalization either in clinical wards or intensive care units, and eventually recovered from severe COVID-19. Sera from these individuals were obtained at a median of 105 days (range, 12-147) after molecular diagnosis of SARS-CoV-2 infection.

Group 4 (control group) included 10 healthy unvaccinated individuals (median age 52 years; range, 35-60 years), with no evidence of prior SARS-CoV-2 infection.

The study was approved by INCLIVA Research Ethics Committee. Informed consent was obtained from all participants.

### Generation of SARS-CoV-2 with variant spikes

We introduced mutations in a mammalian expression vector encoding a codon-optimized SARS-CoV-2 S sequence from the Wuhan reference strain [14]. First, the D614G mutation was introduced by site-directed mutagenesis. Subsequently, additional site-directed mutagenesis and/or the cloning of synthetic fragments harboring the mutations (Gblocks, IDT) was performed to introduce all mutations using the NEBuilder HiFi DNA assembly mix (NEB). For the Beta variant (PANGO Lineage: B.1.351; mutations D80A, L241Del, L242Del, A243Del, E484K, N501Y, D614G, and A701V), a synthetic fragment encoding mutations D80A, L241Del, L242Del, A243Del, E484K, and N501Y was cloned, and the A701V mutation introduced by site-directed mutagenesis. For the Gamma variant (PANGO Lineage: P1 or B.1.1.28.1; mutations L18F, T20N, P26S, D138Y, R190S, K417T, E484K, N501Y, D614G, H655Y, T1027I, and V1176F), a synthetic fragment encoding mutations (L18F, T20N, P26S, D138Y, R190S, K417T, E484K, and N501Y) was cloned, and additional mutations H655Y, T1027I, and V1176F were inserted by site-directed mutagenesis. For the Delta variant (PANGO lineage: B.1.617.2; mutations T19R, del157/158, L452R, T478K, D614G, P681R, and D950N) mutations were introduced by site-directed mutagenesis. Finally, for the Epsilon variant (PANGO lineages B.1.427/B.1.429; mutations S13I, W152C, L452R, D614G), a synthetic fragment encoding mutations S13I, W152C, and L452R was cloned. All plasmids were verified by Sanger sequencing.

### Virus neutralization assay

The neutralization capacity of circulating antibodies (NtAb) against the SARS-CoV-2 S protein was assessed using a GFP-expressing vesicular stomatitis virus pseudotyped with different Spike variants as previously described [15], but using A549-ACE2-TMPRSS2 cells (InvivoGen catalog code a549-hace2tps). All tests were done in duplicate using 5-fold serum dilutions ranging from 1:20 to 1:62,500, with ∼1,000 focus forming units per well. Following 16 hours of infection, the GFP signal in each well was quantified using a live-cell microscope (Incucyte S3, Sartorius). Background fluorescence from uninfected wells was subtracted from all infected well, and the GFP fluorescence in each antibody-treated dilution was standardized to the average fluorescence observed in mock-treated wells. Any value resulting in a relative GFP signal of <0.001 versus was assigned a value of 0.001 to eliminate negative values. Finally, the reciprocal antibody dilution resulting in 50% virus neutralization was calculated using the drc package (version 3.0□1) in R via a three□parameter log□logistic regression model (LL.3 model). For the Beta, Gamma, and Epsilon variants, all assays for the same serum were performed simultaneously with the Wuhan reference strain unless neutralization was repeated due to high variability. As the Delta variant was performed subsequently, we retested each serum was against both Delta and the Wuhan strains unless no neutralization of Wuhan was observed in the previous assays. Sera testing negative (undetectable) were arbitrarily ascribed a titer of 1/20.

### Quantitation of SARS-CoV-2 receptor binding domain-reactive antibodies

Total antibodies (IgG and IgM) against the SARS-CoV-2 S protein receptor-binding domain (RBD) were measured by Roche Elecsys® electrochemiluminescence sandwich immunoassays (Roche Diagnostics, Pleasanton, CA, USA). The limit of detection of the assay is 0.4 IU/ml and its quantification range is between 0.8 and 250 IU/ml. Plasma specimens were further diluted (1/10) for antibody quantitation when appropriate. The assay is calibrated with the first WHO International Standard and Reference Panel for anti-SARS-CoV-2 antibody [16]. Cryopreserved plasma (−20 °C) specimens were thawed and assayed in singlets. Plasma specimens were diluted (1/10) for antibody quantitation when appropriate.

### Statistical methods

Geometric mean titers (GMTs) are reported throughout the study. Comparison of GMTs and medians across groups was carried out by the Student’s t-test or the Mann-whitney U-test, as appropriate. Correlations between variables of interest were calculated by the Spearman Rank test. Frequency comparison across groups was performed using Fisher’s exact test. Two-sided exact P values were reported. A P value <0.05 was considered statistically significant. The analyses were performed using SPSS version 20.0 (SPSS, Chicago, IL, USA).

## RESULTS

### Detection of SARS-CoV-2-RBD and neutralizing antibodies in participants and controls

Antibodies targeting the SARS-CoV-2 RBD were detectable in all but one (belonging to group 3) out of the 67 participants and in none of the controls. Antibody levels across the study groups, shown in Figure 1, were found to differ significantly, with nursing home residents displaying higher levels as compared to that in participants from the other two study groups. Similarly, using a pseudotyped vesicular stomatitis virus system, NtAb against the ancestral strain were detected in all but one (belonging group 3) participants and in none of the controls. Measurable NtAb against the different SARS-CoV-2 VOCs included in the study were found at variable frequencies depending upon the study group and the virus variant considered (Table 1). In all groups, some participants lacked NtAb against one or more VOCs, most notably the Beta variant (15-20%). As for vaccinated individuals (groups 1 and 2), no significant differences in the detection rate of NtAb for any SARS-CoV-2 VOCs were found (*P*≥0.5) (Table 1).

**Table 1.** Detectable neutralizing antibodies against different SARS-CoV-2 variants across study groups.

| SARS-CoV-2 variant | Number of participants with detectable neutralizing antibodies /total participants (%)<sup>a</sup> |  |  |
| --- | --- | --- | --- |
|  | Group 1 | Group 2 | Group 3 |
| Ancestral (Wuhan-Hu-1) | 30/30 (100) | 18/18 (100) | 18/19 (94.7) |
| Beta | 24/30 (80) | 16/18 (88.9) | 16/19 (84.2) |
| Gamma | 27/30 (90) | 17/18 (94.4) | 17/19 (89.5) |
| Delta | 28/30 (93.3) | 18/18 (100) | 17/19 (89.5) |
| Epsilon | 28/30 (93.3) | 17/18 (94.4) | 17/19 (89.5) |
| <sup>a</sup>Group 1, nursing home residents fully vaccinated with the Comirnaty® vaccine; Group 2, healthy individuals fully vaccinated with the Comirnaty® vaccine; Group 3, unvaccinated, COVID-19 recovered individuals that required hospitalization in internal medicine or intensive care units. |  |  |  |

**Figure 1.**
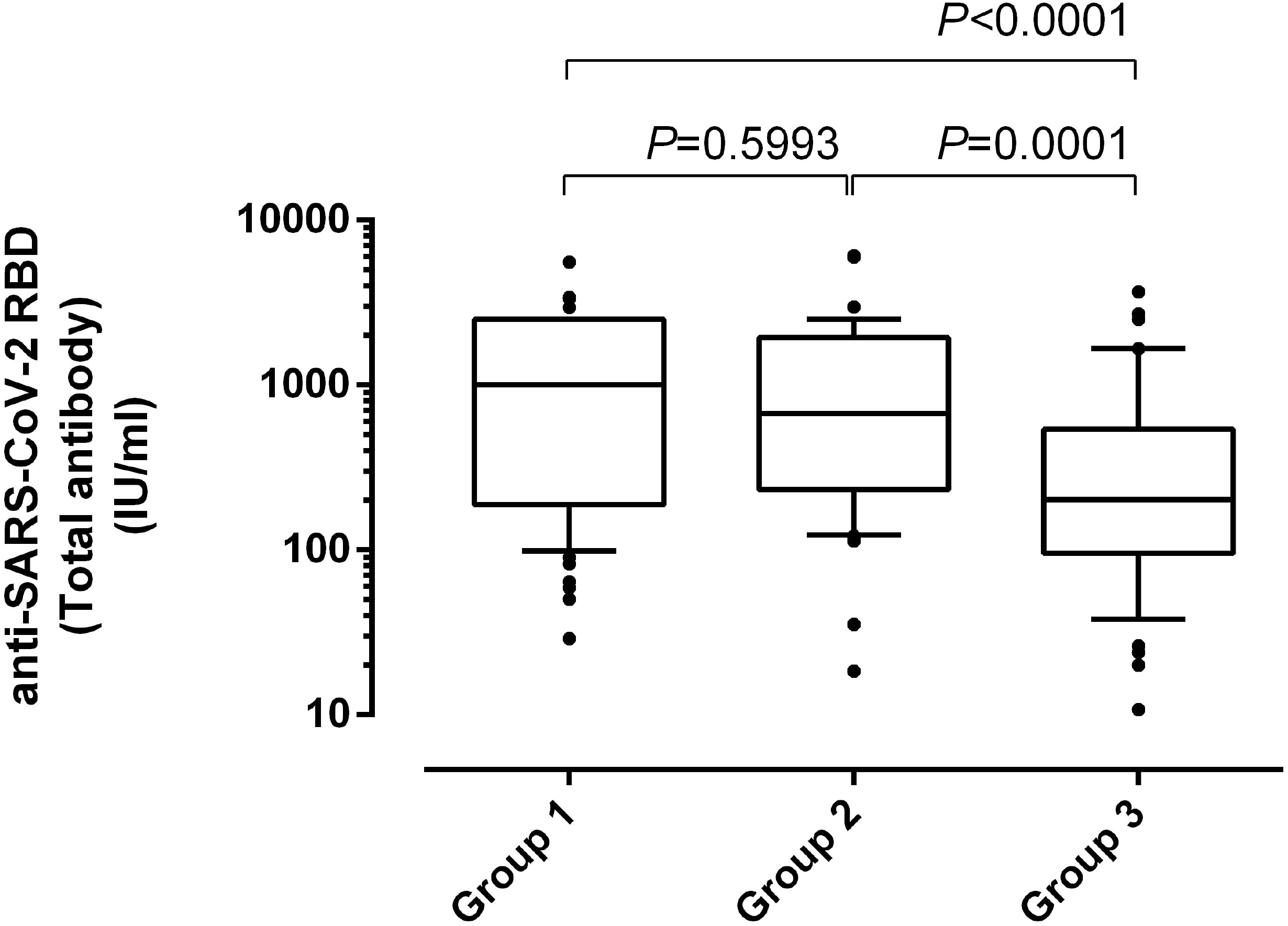
Whisker plots depicting initial anti-SARS-CoV-2 RBD total antibody levels, as measured by the Roche Elecsys® electrochemiluminescence sandwich immunoassays (Roche Diagnostics, Pleasanton, CA, USA), in nursing home residents (Group 1), healthy individuals fully vaccinated with the Comirnaty® vaccine (Group 2) and unvaccinated, COVID-19 recovered individuals that required hospitalization in internal medicine or intensive care units (Group 3). *P* value for comparison across the three groups is shown.

### Antibody neutralizing titers against SARS-CoV-2 S variants across study groups

Overall, all VOCs were neutralized to a lesser extent than the ancestral strain (Figure 2). Specifically, serum NtAb titers were lowest against the Beta variant followed by Gamma, Epsilon, and Delta variants, although wide ranges were observed for all variants. This pattern was shared by all study groups (Figure 3 and Table 2). Accordingly, fold change reduction in NtAb titers relative to the ancestral strain was greatest for the Beta variant (6.7-18.8) followed by Gamma (3.6-6.2), Epsilon (2.9-5.8), and Delta (3.5-4.3) variants. The decrease in NtAb titers for any of SARS-CoV-2 VOCs was not significantly different across comparison groups (Supplementary Figure 1).

**Figure 2.**
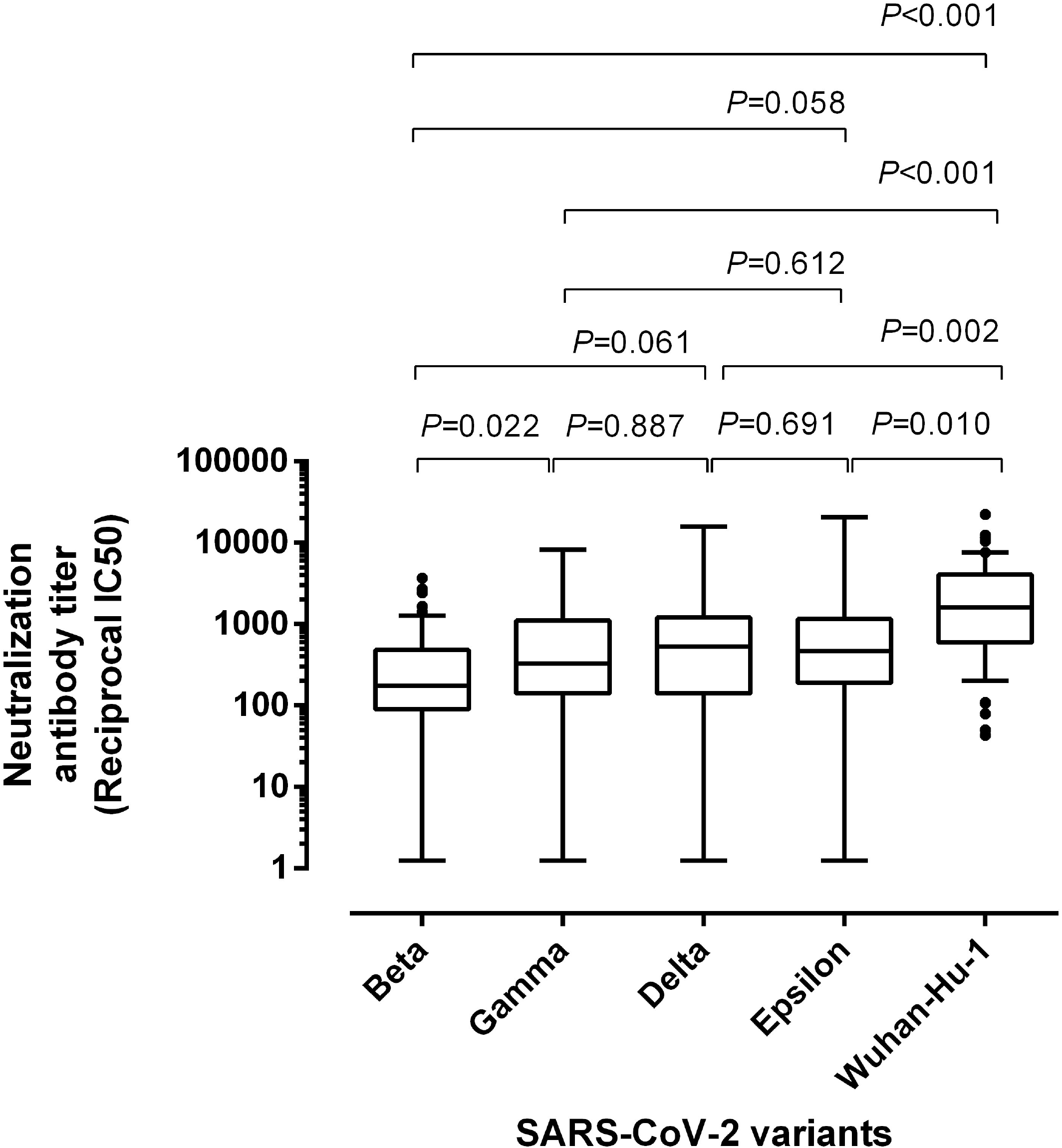
Neutralizing antibody titers against the SARS-CoV-2 ancestral strain (Wuhan-Hu-1) and variants of concern Beta, Gamma, Delta and Epsilon in sera form participants. P values for pairwise comparisons are shown.

**Figure 3.**
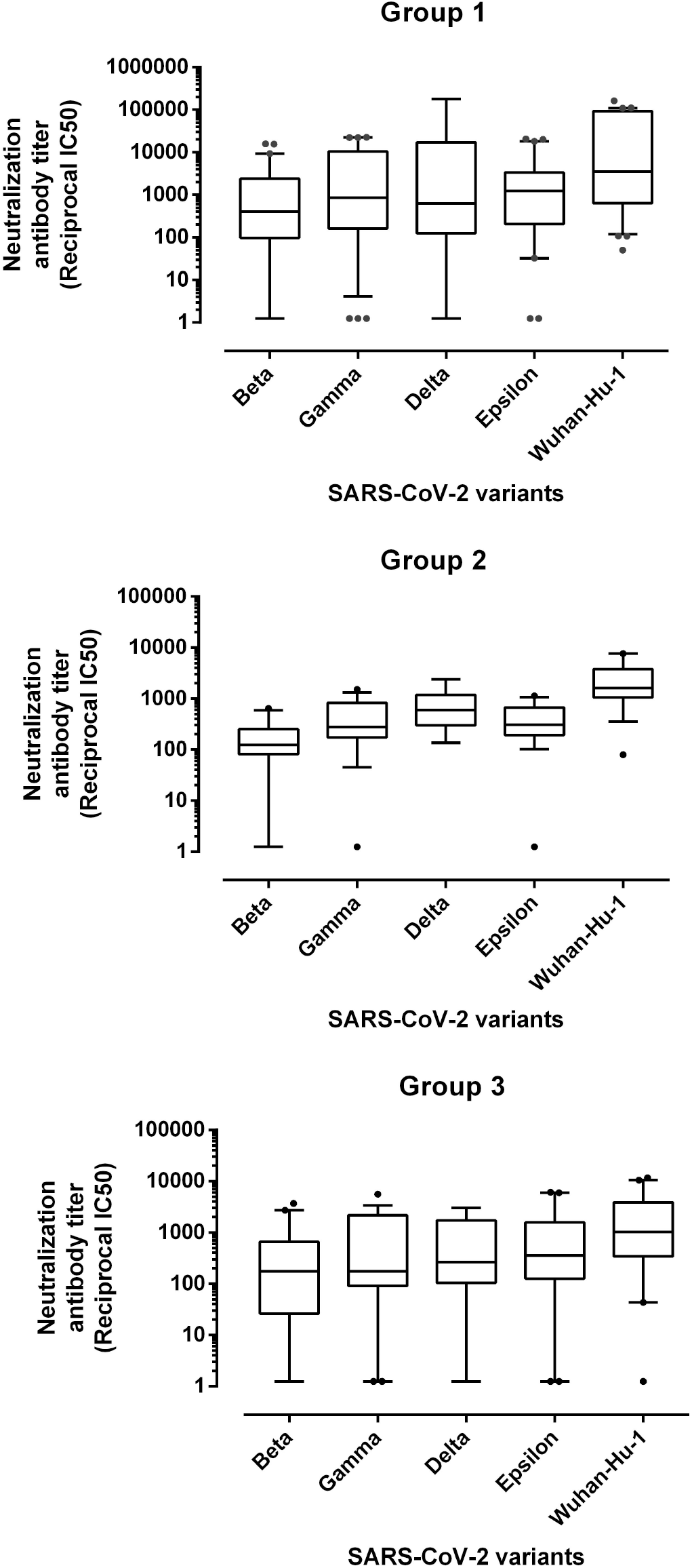
Neutralizing antibody titers against the SARS-CoV-2 ancestral strain (Wuhan-Hu-1) and variants of concern Beta, Gamma, Delta and Epsilon across study groups: nursing home residents (Group 1), healthy individuals fully vaccinated with the Comirnaty® vaccine (Group 2) and unvaccinated, COVID-19 recovered individuals that required hospitalization in internal medicine or intensive care units (Group 3).

**Figure 4.**
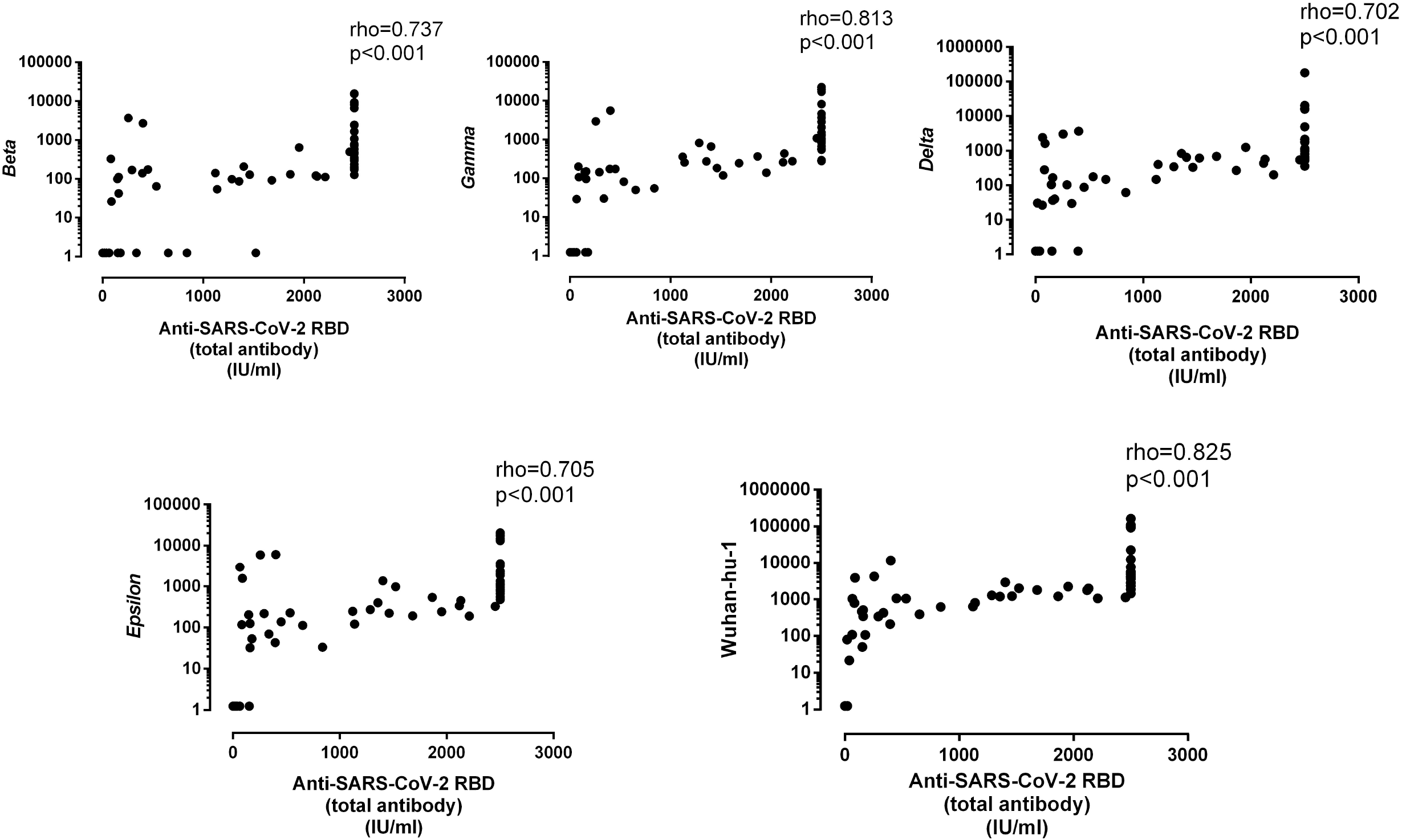
Overall correlation between neutralizing antibody titers and anti-SARS-CoV-2 RBD total antibody levels, as measured by the Roche Elecsys® electrochemiluminescence sandwich immunoassays (Roche Diagnostics, Pleasanton, CA, USA), according to the SARS-CoV-2 S-variant. Rho and *P* values are shown.

**Table 2.** Serum Neutralizing antibody titers against SARS-CoV-2 variants across study groups.

| SARS-CoV-2 variant | Parameter | Study groups<sup>a</sup> |  |  |
| --- | --- | --- | --- | --- |
|  |  | Group 1 | Group 2 | Group 3 |
| Beta | GMT (SD) | 238 (4,510) | 124 (316) | 122 (999) |
|  | Mean fold change reduction of NtAb titer (CI 95%)<sup>b</sup> | 18.8 (109.1) | 13.1 (6.9) | 6.7 (84.9) |
| Gamma | GMT (SD) | 722 (8,612) | 256 (435) | 225 (1607) |
|  | Mean fold change reduction of NtAb titer (CI 95%)<sup>b</sup> | 6.2 (9.1) | 6.3 (6.6) | 3.6 (4.8) |
| Delta | GMT (SD) | 1,037 (67,237) | 543 (1,138) | 231 (1,162) |
|  | Mean fold change reduction of NtAb titer (CI 95%)<sup>b</sup> | 4.3 (10.9) | 3.0 (0.6) | 3.5 (4.1) |
| Epsilon | GMT (SD) | 774 (6,778) | 261 (324) | 283 (1,869) |
|  | Mean fold change reduction of NtAb titer (CI 95%)<sup>b</sup> | 5.8 (7.7) | 6.2 (6.6) | 2.9 (2.2) |
| (Wuhan-Hu-1) | GMT (SD) | 4,509 (48,923) | 1,622 (2,259) | 442 (2,083) |
| CI, confidence interval; GMT, geometric mean titer; NtAb, neutralizing antibodies; SD, standard deviation.<br><sup>a</sup>Group 1, nursing home residents fully vaccinated with the Comirnaty® vaccine; Group 2, healthy individuals fully vaccinated with the Comirnaty® vaccine; Group 3, unvaccinated, COVID-19 recovered individuals that required hospitalization in internal medicine or intensive care units.<br><sup>b</sup>Relative to the NtAb titer against WA1/2020. |  |  |  |  |

### Correlation between SARS-CoV-2 neutralizing and anti-RBD antibodies

The degree of correlation between levels of SARS-CoV-2 antibodies neutralizing SARS-CoV-2 VOCs and total anti-RBD antibodies across the different population groups was examined next. Overall, the correlation between these two parameters was best for the ancestral strain and slightly decreased for all VOCs (Supplementary Figure 1). Of interest, this pattern was similar for both the vaccinated participants from nursing home residents and healthy controls (Supplementary Table 1).

## DISCUSSION

Reduced neutralization of VOCs as compared to the ancestral Wuhan-Hu-1 strain has been observed in sera from individuals that were fully vaccinated against SARS-CoV-2 or have recovered from infection by either the ancestral Wuhan or the Wuhan/D164G SARS-CoV-2 strains [5-12,17-18]. This has been observed in both the full SARS-CoV-2 virus or pseudotyped virus systems and is most notable for the Beta variant. As for recipients of the Comrinaty® COVID-19 vaccine, this observation has been mostly made in vaccine clinical trials participants and seemingly healthy health workers [5-12]. Here, using S-pseudotyped SARS-CoV-2 variants, we extend this finding to include elderly nursing home residents, both in SARS-CoV-2 naïve participants and those infected by the Wuhan/D614G variant prior to vaccination. In effect, one month after the second vaccine dose, sera from fully vaccinated nursing home residents retained the ability to neutralize VOCs in most cases, but did so to a lesser extent than the ancestral strain, especially in the case of the Beta variant. Importantly, reduction in NtAb activity against VOCs in this population group was not significantly different from that seen in healthy fully-vaccinated controls (in sera obtained a median of two weeks after vaccination) and unvaccinated individuals who had recovered from severe COVID-19 diagnosed within a median time of 3 months. Leaving aside participants who remained unvaccinated and naturally contracted the infection, which are not strictly comparable to vaccinees, our data strongly support the idea that older age, frailty, and concurrence of co-morbidities in fully-vaccinated individuals had no impact on the serum NtAb activity profile against VOCs, including Delta variant that currently dominates in many countries. The reduction in NtAb activity for all VOCs relative to that observed for the Wuhan-Hu-1 reference strain (fold-change reduction in NtAb activity) was higher for all VOCs compared to other studies, especially the Beta variant. For example, using a pseudotyped virus system, Liu et al. [5-7] reported a 2.7- and 1.4-fold reduction in the NtAb activity against Beta and Delta variants, respectively, for sera from participants in the Comirnaty® COVID-19 vaccine the trial, who were seemingly comparable to our healthy vaccinated controls (13.1 and 3.0 fold reduction, respectively). In turn, using live virus isolates in microneutralization assays, Lustig and colleagues [11] found fold change reduction in NtAb titers of 10.4, 2.3, and 2.1 against the Beta, Gamma, and Delta variants compared with the ancestral virus in health care workers fully vaccinated with the Comirnaty® COVID-19 vaccine. Comparison of fold change reduction figures across studies is not straightforward, due not only to differences between population group characteristics, especially in terms of age, sex, comorbidities, and SARS-CoV-2 infection status prior to vaccination, but also to differences in the pseudotyped virus platforms, the chosen S sequences used to represent each variant or the use of live viruses. Despite these differences, a similar pattern of NtAb activity against VOCs can be derived from these studies as well as our current work, with the Beta variant being less efficiently neutralized than the ancestral strain, followed by the Gamma, Epsilon, and Delta variants.

Levels of total antibodies binding SARS-CoV-2 RBD as quantitated by the Roche Elecsys® electrochemiluminescence sandwich immunoassays strongly correlate with NtAb titers against the ancestral SARS-CoV-2 strain [19,20]. Here we extended this observation to those against Beta, Gamma, Delta, and Epsilon variants, suggesting that SARS-CoV-2 RBD antibody levels measured by this assay may be a reliable proxy for serum NtAb activity against these VOCs, regardless of the vaccinated population group considered.

The relatively small sample size could be considered a limitation of the current study. Also, a limitation inherent to the use of S-pseudotyped viruses is the inability to assess the impact of variations in other viral genes outside of the S protein that could affect sera neutralization.

In summary, herein we show that neutralizing activity of sera against SARS-CoV-2 variants carrying critical escape mutations in the S gene is decreased relative to the ancestral strain in both qualitative and quantitative terms in nursing home residents recently vaccinated with the Comirnaty® COVID-19 vaccine. Nevertheless, the fold reduction in NtAb activity in this population group was not different from that seen in vaccinated controls and recovered COVID-19 individuals who had not been vaccinated. Importantly, among VOCs, the Delta variant that is currently predominant worldwide was best neutralized by sera from participants in all study groups. Nevertheless, the degree to which reduced neutralization activity of the different VOCs, and especially the Delta variant, impact infection and/or disease in vaccinated nursing home residents translate into diminished effective protection is currently under investigation.

## Data Availability

The data that support the findings of this study are available from the corresponding author, (DN), upon reasonable request.

## ACKNOWLEDGMENTS

We are grateful to all personnel who work at nursing home residences affiliated with the Clínico-Malvarrosa Health Department and at Clinic University Hospital, in particular to those at the Microbiology laboratory, for their commitment in the fight against COVID-19. Ignacio Torres holds a Río Hortega Contract (CM20/00090) from the Carlos III Health Institute. Eliseo Albert holds a Juan Rodés Contract (JR20/00011) from the Carlos III Health Institute. Estela Giménez holds a Juan Rodés Contract (JR18/00053) from the Carlos III Health Institute. Ron Geller holds a Ramon y Cajal Fellowship from the Spanish Ministry of Economy and Competitiveness (RYC-2015-17517).

## AUTHOR CONTRIBUTIONS

BS-S, EA, JZ, IT, and EG: Methodology and data validation. PB, MJB, CR: in charge of implementing public health policies to combat SARS-CoV-2 epidemic at nursing home residences affiliated to the Clínico-Malvarrosa Health Department. RG and DN: Conceptualization, supervision, writing the original draft. All authors reviewed the original draft.

## FIGURE LEGENDS

**Supplementary Figure 1**. Fold change reduction in neutralizing antibody titers against SARS-CoV-2 variants of concern as compared to the ancestral strain as a function of the study group considered: nursing home residents (Group 1), healthy individuals fully vaccinated with the Comirnaty® vaccine (Group 2) and unvaccinated, COVID-19 recovered individuals that required hospitalization in internal medicine or intensive care units (Group 3).

