## Supplementary figures and images for "Neutralizing antibodies against SARS-CoV-2 variants of concern elicited by the Comirnaty® COVID-19 vaccine in nursing home residents"

### Supplementary Figure 1

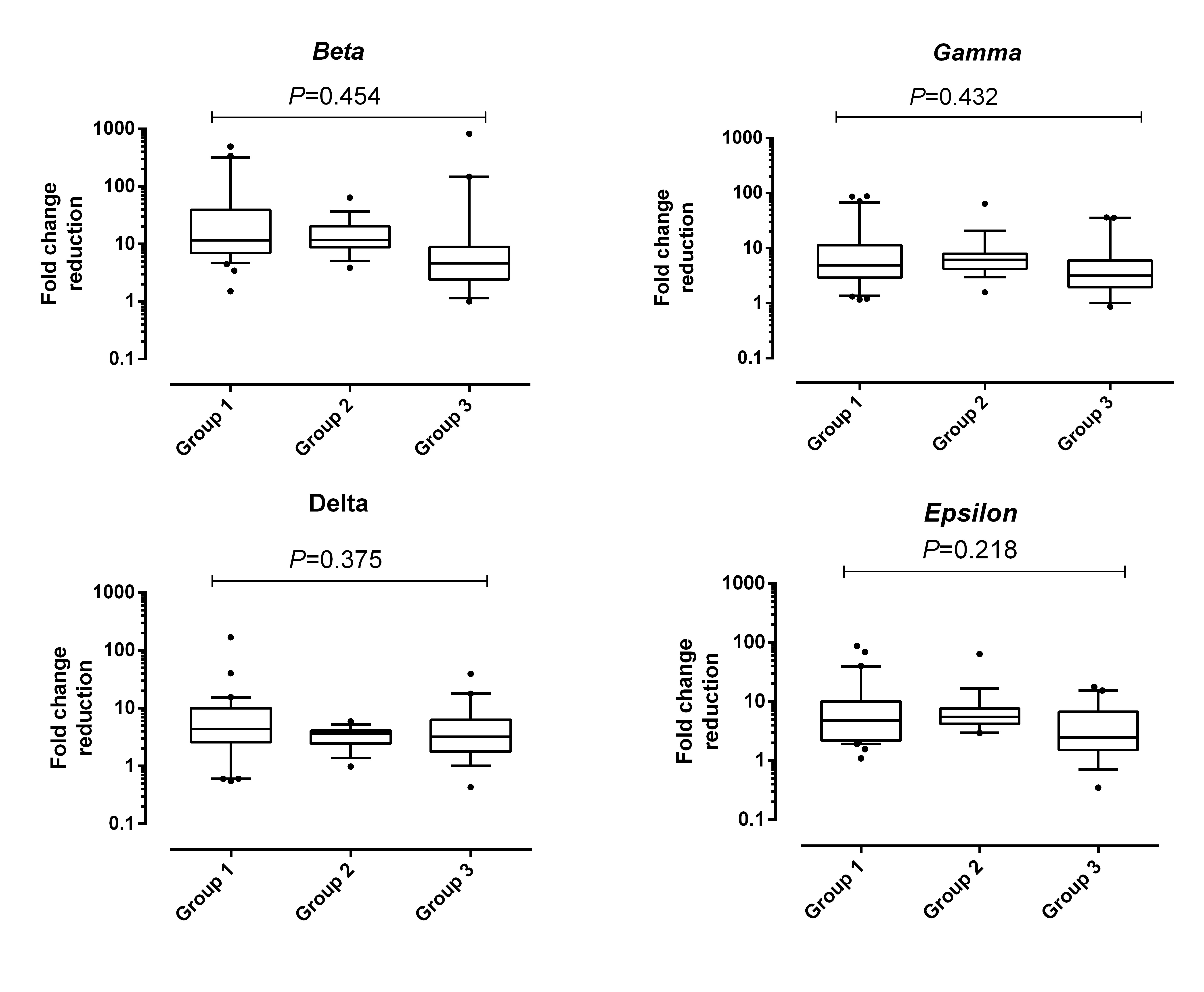
