## Supplementary Table 1 for "Neutralizing antibodies against SARS-CoV-2 variants of concern elicited by the Comirnaty® COVID-19 vaccine in nursing home residents"

| **Supplementary Table 1. Correlation between SARS-CoV-2 neutralizing antibodies and anti-RBD antibodies in fully vaccinated nursing home residents and healthy controls.** | | | | |
| --- | --- | --- | --- | --- |
| SARS-CoV-2 variant | Group 1^a^ | | Group 2^b^ | |
|  | Rho value^c^ | *P value* | Rho value^c^ | *P value* |
| Beta | 0.84 | <0.001 | 0.81 | <0.001 |
| Gamma | 0.83 | <0.001 | 0.83 | <0.001 |
| Delta | 0.81 | <0.001 | 0.79 | <0.001 |
| Epsilon | 0.82 | <0.001 | 0.85 | <0.001 |
| Wuhan-Hu-1 | 0.84 | <0.001 | 0.88 | <0.001 |
| ^a^Nursing home residents and ^b^Healthy individuals fully vaccinated with the Comirnaty® vaccine.  ^c^Spearman Rank test | | | | |
